# Early estimates of SARS-CoV-2 Omicron variant severity based on a matched cohort study, Ontario, Canada

**DOI:** 10.1101/2021.12.24.21268382

**Authors:** Ana Cecilia Ulloa, Sarah A. Buchan, Nick Daneman, Kevin A Brown

**Affiliations:** Public Health Ontario, Toronto, Canada (Ulloa, Buchan, Daneman, Brown)

## Abstract

While it is now evident that Omicron is rapidly replacing Delta, largely due to immune escape, it is less clear how the severity of Omicron compares to Delta. In Ontario, we sought to examine hospitalization and death associated with Omicron, as compared to cases infected with Delta. We conducted a matched cohort study, considering time to hospitalization or death as the outcome. Cases were matched on gender, age, vaccination status, health region and onset date. We identified 29,594 Omicron cases that met eligibility criteria, of which 11,622 could be matched with at least one Delta case (N=14,181). There were 59 (0.51%) hospitalizations and 3 (0.03%) deaths among matched Omicron cases, compared to 221 (1.6%) hospitalizations and 17 (0.12%) deaths among matched Delta cases. The risk of hospitalization or death was 65% lower (hazard ratio, HR=0.35, 95%CI: 0.26, 0.46) among Omicron cases compared to Delta cases, while risk of intensive care unit admission or death was 83% lower (HR=0.17, 95%CI: 0.08, 0.37). While severity is likely to be reduced, the absolute number of hospitalizations and impact on the healthcare system may nevertheless be significant due to the increased transmissibility of Omicron.

## Introduction

The WHO designated Omicron as a Variant of Concern on November 26, 2021^1^. Omicron has more than 30 mutations in the spike protein, and it is now evident that Omicron is rapidly replacing Delta^2^ due to increased levels of immune escape. It is less clear how the severity of Omicron compares to Delta. Early data from South Africa suggests that Omicron may be less severe;^3^ however, the low average age, extent of previous infection, and low vaccination rates impact generalizability to other countries. In Ontario, we sought to examine rates of hospitalization, intensive care unit admission, and death associated with Omicron, as compared to matched patients infected with Delta.

## Methods

We conducted a retrospective population-wide matched cohort study of patients infected with Omicron and Delta variants, using Ontario’s Public Health Case and Contact Management Solution (CCM), a database containing all diagnosed SARS-CoV-2 infections, linked to Ontario Ministry of Health’s COVaxON application, containing all COVID-19 vaccination records in the province. Cases were included if onset occurred between November 22 (first Omicron case in Ontario) and December 25, 2021. Case onset date was defined by symptom onset, or for asymptomatic cases, specimen collection. Cases were excluded if they were missing onset date, age, gender, or were hospital-acquired. Omicron cases included those identified by whole genome sequencing (WGS), S-gene target failure (SGTF) with cycle threshold ≤30 before December 13 (date of 50% Omicron prevalence)^2^, and any SGTF on or after December 13. Delta cases included those detected by WGS, those with amplification of the S-gene, and all cases not identified as Omicron prior to December 3 (date of 5% Omicron prevalence)^2^. Omicron cases were matched to Delta cases using up to 2:1 matching on gender (male, female, other), age (0-4, 5-11, 12-18, 19-29, 30-39, 40-49, 50-59, 60-69, 70-79, 80+), vaccination status (0 doses, 1 dose, 2 doses, 3 doses), time since most recent dose (>7 or 14 days to <3 months, 3-6 months, >6 months), region and onset date (±7 days). Cox proportional hazards models accounting for clustering within matched sets were used to determine the hazards of hospitalization or death, and intensive care unit (ICU) admission, compared to Delta cases. Due to concerns around incidental SARS-CoV-2 findings upon hospital admission due to universal screening practices in hospitals, we conducted a separate sensitivity analysis excluding cases with first positive specimen collection on the day of or the day prior to hospitalization. Stratified hazards were estimated by gender, age group, and vaccination status.

## Results

We identified 29,594 Omicron cases that met eligibility criteria, of which 11,622 were matched with ≥1 Delta case (N=14,181, Table 1). There were 59 (0.51%) hospitalizations and 3 (0.03%) deaths among matched Omicron cases, compared to 221 (1.56%) hospitalizations and 17 (0.12%) deaths among matched Delta cases. The risk of hospitalization or death was 65% lower (hazard ratio, HR=0.35, 95%CI: 0.26, 0.46, Table 2) in Omicron compared to Delta cases (Figure 1), and the risk of ICU admission or death was 83% lower (HR=0.17, 95%CI: 0.08, 0.37). A sensitivity analysis excluding cases with potential incidental SARS-CoV-2 findings on hospital admission, also demonstrated reduced severity of Omicron relative to Delta (HR=0.25, 95%CI: 0.16, 0.41). Stratified estimates of Omicron severity by age, gender, and vaccination status all indicated reduced severity of Omicron (Table 2).

**Table 1.** Demographic characteristics, vaccination status, and outcomes among SARS-CoV-2 Delta and Omicron variant cases, and among matched cases (N, %).

|  | Full Cohort |  | Matched Cohort |  |
| --- | --- | --- | --- | --- |
|  | Delta | Omicron | Delta | Omicron |
|  | N=22,769 | N=29,594 | N=14,181 | N=11,622 |
| Age (median, [IQR]) | 33.0 [12.0, 49.0] | 30.0 [21.0, 44.0] | 31.0 [14.0, 46.0] | 31.0 [16.0, 46.0] |
| Gender |  |  |  |  |
| Female | 11,198 (49.2) | 14,779 (49.9) | 7,062 (49.8) | 5,783 (49.8) |
| Male | 11,513 (50.6) | 14,783 (50.0) | 7,107 (50.1) | 5,831 (50.2) |
| Other | 58 (0.3) | 32 (0.1) | 12 (0.1) | 8 (0.1) |
| Vaccination status (doses, and time since last dose) |  |  |  |  |
| 0 doses | 10,286 (45.2) | 4,005 (13.5) | 5,108 (36.0) | 3,629 (31.2) |
| 1 dose |  |  |  |  |
| >14d to <3 months | 1037 (4.6) | 1,155 (3.9) | 827 (5.8) | 724 (6.2) |
| 3 to 6 months | 139 (0.6) | 87 (0.3) | 59 (0.4) | 46 (0.4) |
| >6 months | 133 (0.6) | 102 (0.3) | 46 (0.3) | 39 (0.3) |
| 2 doses |  |  |  |  |
| >7d to <3 months | 616 (2.7) | 1,025 (3.5) | 381 (2.7) | 320 (2.8) |
| 3 to 6 months | 5,442 (23.9) | 12,958 (43.8) | 4,166 (29.4) | 3,719 (32.0) |
| >6 months | 4,348 (19.1) | 8,469 (28.6) | 3,125 (22.0) | 2,731 (23.5) |
| 3 doses |  |  |  |  |
| >7d to <3 months | 721 (3.2) | 1,710 (5.8) | 450 (3.2) | 396 (3.4) |
| 3 to 6 months | 47 (0.2) | 83 (0.3) | 19 (0.1) | 18 (0.2) |
| Outcomes |  |  |  |  |
| Hospitalization/death | 601 (2.64) | 75 (0.25) | 221 (1.56) | 59 (0.51) |
| ICU admission/death | 190 (0.83) | 8 (0.03) | 60 (0.42) | 7 (0.06) |
| Deaths | 83 (0.36) | 3 (0.01) | 17 (0.12) | 3 (0.03) |

Note: d, days; IQR, inter-quartile range; ICU, intensive care unit

**Table 2:** Risk of hospitalization, intensive care unit admission, and death associated with Omicron cases relative to matched Delta cases, Ontario, Canada. All analyses are based on proportional hazards models and presented as hazards ratios for Omicron relative to Delta. Hazards ratios of 1 indicate equal risk for Omicron relative to Delta, <1 indicate reduced severity of Omicron, >1 indicate increased severity of Omicron.

|  | Delta cases<br>N (%) | Omicron cases<br>N (%) | Hospitalization or death,<br>HR (95%CI) | ICU admission or death,<br>HR (95%CI) | Hospitalization or death SA*,<br>HR (95%CI) |
| --- | --- | --- | --- | --- | --- |
| Total | 14,181 (100) | 11,622 (100) | 0.35 (0.26, 0.46) | 0.17 (0.08, 0.37) | 0.25 (0.16, 0.41) |
| Stratified analyses |  |  |  |  |  |
| Gender |  |  |  |  |  |
| Female | 7,062 (49.8) | 5,783 (49.8) | 0.36 (0.25, 0.54) | 0.24 (0.08, 0.68) | 0.29 (0.15, 0.54) |
| Male | 7,107 (50.1) | 5,831 (50.2) | 0.33 (0.23, 0.49) | 0.12 (0.04, 0.39) | 0.22 (0.10, 0.46) |
| Age |  |  |  |  |  |
| <60 | 12,754 (89.9) | 10,482 (90.2) | 0.30 (0.19, 0.48) | 0.05 (0.01, 0.38) | 0.30 (0.14, 0.63) |
| ≥60 | 1,427 (10.1) | 1,140 (9.8) | 0.40 (0.28, 0.56) | 0.28 (0.11, 0.67) | 0.24 (0.13, 0.46) |
| Vaccine doses |  |  |  |  |  |
| 0 doses | 5,108 (36.0) | 3,629 (31.2) | 0.40 (0.28, 0.56) | 0.16 (0.05, 0.50) | 0.17 (0.06, 0.46) |
| 2 doses | 7,672 (54.1) | 6,770 (58.3) | 0.40 (0.27, 0.58) | 0.12 (0.03, 0.51) | 0.25 (0.13, 0.50) |

CI=confidence interval; HR=hazard ratio; ICU=intensive care unit; \*sensitivity analysis excludes Delta or Omicron cases with potential incidental SARS-CoV-2 findings (first positive specimen collection on the day of or the day prior to hospitalization)

**Figure 1.**
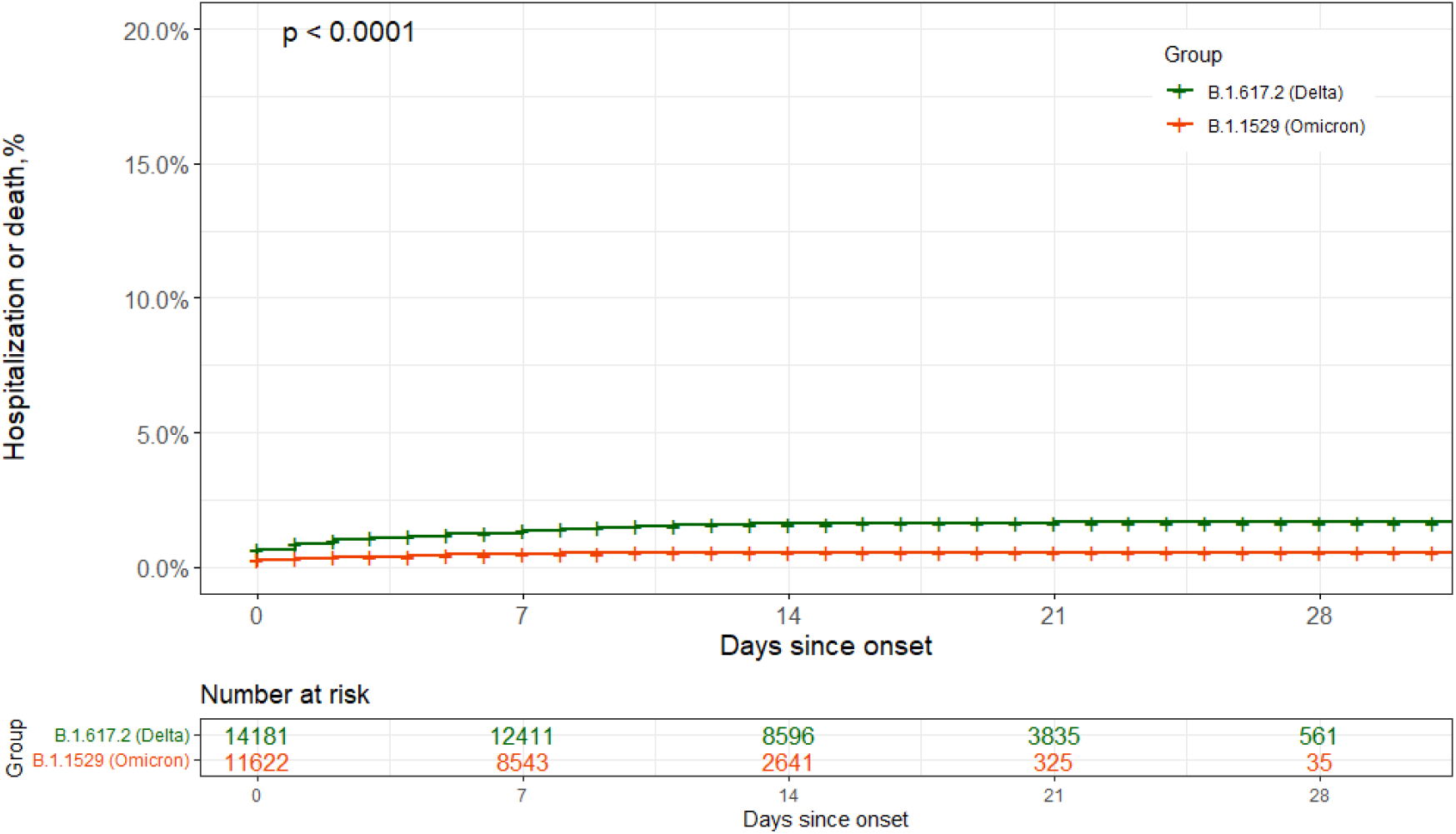
SARS-CoV-2 associated hospitalization or death among matched Omicron variant (N=11,622, orange line) and Delta variant cases (N=14,181, green line) cases as a function of days since onset. Date of onset was defined as symptom onset date, falling back to first positive specimen collection date in the absence of a documented symptom onset date.

## Discussion

In this matched study of over 11,000 Omicron cases in Ontario, Canada, we found that the risk of hospitalization was 65% lower for SARS-CoV-2 cases infected with the Omicron variant relative to the Delta variant. Our results align with findings from South Africa, Scotland and England, all of which have demonstrated substantial decreases in risk of hospitalization associated with Omicron.^3-5^ One early hypothesis for increased transmissibility with decreased severity is the increased replication in the bronchi but decreased replication in lung parenchyma.^6^

Our study has some limitations, in particular the short follow-up duration and potential misclassification due to incidental findings from hospital admission screening, and incomplete public health follow-up as incidence increased. Nevertheless, Omicron appears to demonstrate lower disease severity for both vaccinated and unvaccinated individuals. While severity is likely to be reduced, the absolute number of hospitalizations and impact on the healthcare system is likely to be significant due to the large number of Omicron infections.

## Data Availability

The data from this study is not publically available.

